# CAPoxy: a feasibility study to investigate multispectral imaging in nailfold capillaroscopy

**DOI:** 10.1101/2025.08.01.25332687

**Authors:** Michaela Taylor-Williams, Irsa Khalil, Joanne Manning, Graham Dinsdale, Michael Berks, Luca Porcu, Sarah Wilkinson, Sarah Bohndiek, Andrea Murray

**Author notes:** now at Department of Biomedical Engineering, Johns Hopkins University, USA. Co-corresponding authors: Sarah Bohndiek. Andrea Murray. 01612754292.

## Abstract

**Background:** Nailfold capillaroscopy enables visualisation of structural abnormalities in the microvasculature of patients with systemic sclerosis (SSc). The objective of this feasibility study was to determine whether multispectral imaging could provide functional assessment (differences in haemoglobin concentration or oxygenation) of capillaries to aid discrimination between healthy controls and patients with SSc. MSI of nailfold capillaries visualizes the smallest blood vessels and the impact of SSc on angiogenesis and their deformation, making it suitable for evaluating oxygenation-sensitive imaging techniques. Imaging of the nailfold capillaries offers tissue-specific oxygenation information, unlike pulse oximetry, which measures arterial blood oxygenation as a single-point measurement.

**Methods:** The CAPoxy study was a single-centre, cross-sectional, feasibility study of nailfold capillary multispectral imaging, comparing a cohort of patients with SSc to controls. A nine-band multispectral camera was used to image 22 individuals (10 patients with SSc and 12 controls). Linear mixed-effects models and summary statistics were used to compare the different regions of the nailfold (capillaries, surrounding edges, and outside area) between SSc and controls. A machine learning model was used to compare the two groups.

**Results:** Patients with SSc exhibited higher indicators of haemoglobin concentration in the capillary and adjacent regions compared to controls, which were significant in the regions surrounding the capillaries (p<0.001). There were also spectral differences between the SSc and controls groups that could indicate differences in oxygenation of the capillaries and surrounding tissue. Additionally, a machine learning model distinguished SSc patients from healthy controls with an accuracy of 84%, suggesting potential for multispectral imaging to classify SSc based on structural and functional microvascular changes.

**Conclusions:** Data indicates that multispectral imaging differentiates between patients with SSc from controls based on differences in vascular function. Further work to develop a targeted spectral camera would further improve the contrast between patients with SSc and controls, enabling better imaging.

**Key messages:** Multispectral imaging holds promise for providing functional oxygenation measurement in nailfold capillaroscopy.

Significant oxygenation differences between individuals with systemic sclerosis and healthy controls can be detected with multispectral imaging in the tissue surrounding capillaries.

## Introduction

Blood oxygenation and vascular changes are believed to play an important role in the development of systemic sclerosis (SSc) and Raynaund’s phenomena (RP) (1–4). Patients with SSc experience dysfunction and loss of blood vessels alongside impaired oxygen distribution to the connective tissues, creating a hypoxic environment (5). Vasculopathy and subsequent structural abnormality and dysfunction of the microvasculature result in a cycle of fibrosis of connective tissue and microvascular dysfunction (5,6). The hypoxic environment has been characterised through increase of hypoxia-inducible factor-1 alpha (HIF-1α) (3,5,6), a transcription factor that activates genes essential for oxygen homeostasis (3,5,6).

Furthermore, disruption of vascular endothelial growth factor (VEGF) can lead to the formation of new, yet dysfunctional (angiogenic) blood vessels (7,8). Thus hypoxia appears to play a vital role in the pathogenesis of SSc, by driving a range of tissue abnormalities from vascular disruption to degradation of the extracellular matrix (3,5–8). Similarly, hypoxia is associated with RP, which causes episodic colour changes in fingers on exposure to cold or stress (9). It is caused by vascular spasms with ischaemia (white fingers), hypoxia (blue fingers) and hyperaemia on rewarming (red fingers). Secondary RP is associated with SSc and can be a preceding indicator of SSc (9).

Despite the clear role of hypoxia in the pathogenesis of the disease, due to the difficulty in non-invasive measurements, it has yet to be successfully exploited in the diagnostic pathway. Nailfold capillaroscopy is a non-invasive *in vivo* imaging technique used in the assessment and study of microvascular changes in a range of clinical scenarios, including the diagnosis (10,11) and monitoring of SSc (12). Nailfold capillaroscopy currently relies on purely morphological assessment of structural changes occuring over months to years; the examination magnifies the capillaries for evaluation of the size (width and length), shape, and number of capillaries, either visually, or using software to quantify the changes (11–14). To better characterise SSc, it would be valuable to extend capillaroscopy to examine the functional dynamics of the blood vessels in the nailfold (over timeframes of seconds to hours), which would aid measurement of the physiological drivers underpinning the observed changes in capillary structure (1,15,16).

The absorption of light by haemoglobin in the blood vasculature is the primary source of contrast in traditional nailfold capillaroscopy. Capturing spectral data beyond our colour vision can enable quantitative analysis of the functional tissue response, because the absorption of light by haemoglobin varies according to whether oxygen is bound (oxyhaemoglobin, HbO_2_) or unbound (deoxyhaemoglobin, Hb). Multispectral imaging (MSI), which collects multiple wavelength (colour)-resolved images of a given scene (4,17), could be coupled with capillaroscopy to resolve the haemoglobin content and oxygenation of tissue. Prior work examined a multispectral imaging approach to capillaroscopy in an illumination-based prototype (15), however, due to the slow data acquisition process in that system, it was not suitable to deploy clinically (15). MSI of nailfold capillaries enables direct visualization of the smallest blood vessels in the body and the impact SSc has on angiogenesis and the deformation of these blood vessels, making it a suitable target for evaluating imaging techniques sensitive to oxygenation. Imaging of the nailfold capillaries provides information on tissue-specific oxygenation, opposed to pulse oximetry which measures arterial blood oxygenation and is a single-point measurement.

Here, we present CAPoxy, a feasiblity study to investigate the application of MSI in nailfold capillaroscopy for patients with SSc. The centre wavelength and bandwidth, along with the number of wavelengths examined, define the spectral resolution of the MSI technique (4).

Here, we used a commercial multispectral camera that enabled video-rate imaging of 8 different colour channels, along with a broadband (multiple wavelengths – to collect ‘white light’) response channel. Combining morphological and functional assessment may provide improved sensitivity to subtle changes in disease status, with potential applications in earlier SSc diagnosis or in monitoring response to a therapeutic intervention. In this feasibility study, we took a first step towards assessing how MSI of the nailfold could improve capilliaroscopy with the ultimate goal of informing clinicians about the function of the microvasculature in the nailfold, in addition to the traditional morphological information available from the measurement of microvascular deformation.

The primary purpose of the study was to investigate the application of MSI in nailfold capillaroscopy to discriminate between SSc patients and healthy controls. This included developing a capillaroscopy imaging system to measure, at the individual level, oxygenation and explore differences in oxygenation between the participant groups and how this compares with structural variables.

## Methods and participants

### Imaging system

An MSI capillaroscopy system was designed based on an existing clinical capillaroscopy setup (14,18). The MSI system (Figure 1a) used LED illumination (White Mi-LED Fiber Optic LED Illuminator, Dolan Jenner) coupled to a fibre optic ring light (0.83 inch ID Fiber Optic Ring Light Guide, Edmund Optics) to illuminate the nailfold of the participant. The spectrum of the white LED used for illumination was recorded with a spectrometer (Avantes AvaSpecULS 2048-USB2-FCPC) and the change in irradiance of the white LED with time was calibrated using a power meter (Thorlabs PM400) at 15-second intervals (Supplementary Figure 1a,b).

**Figure 1:**
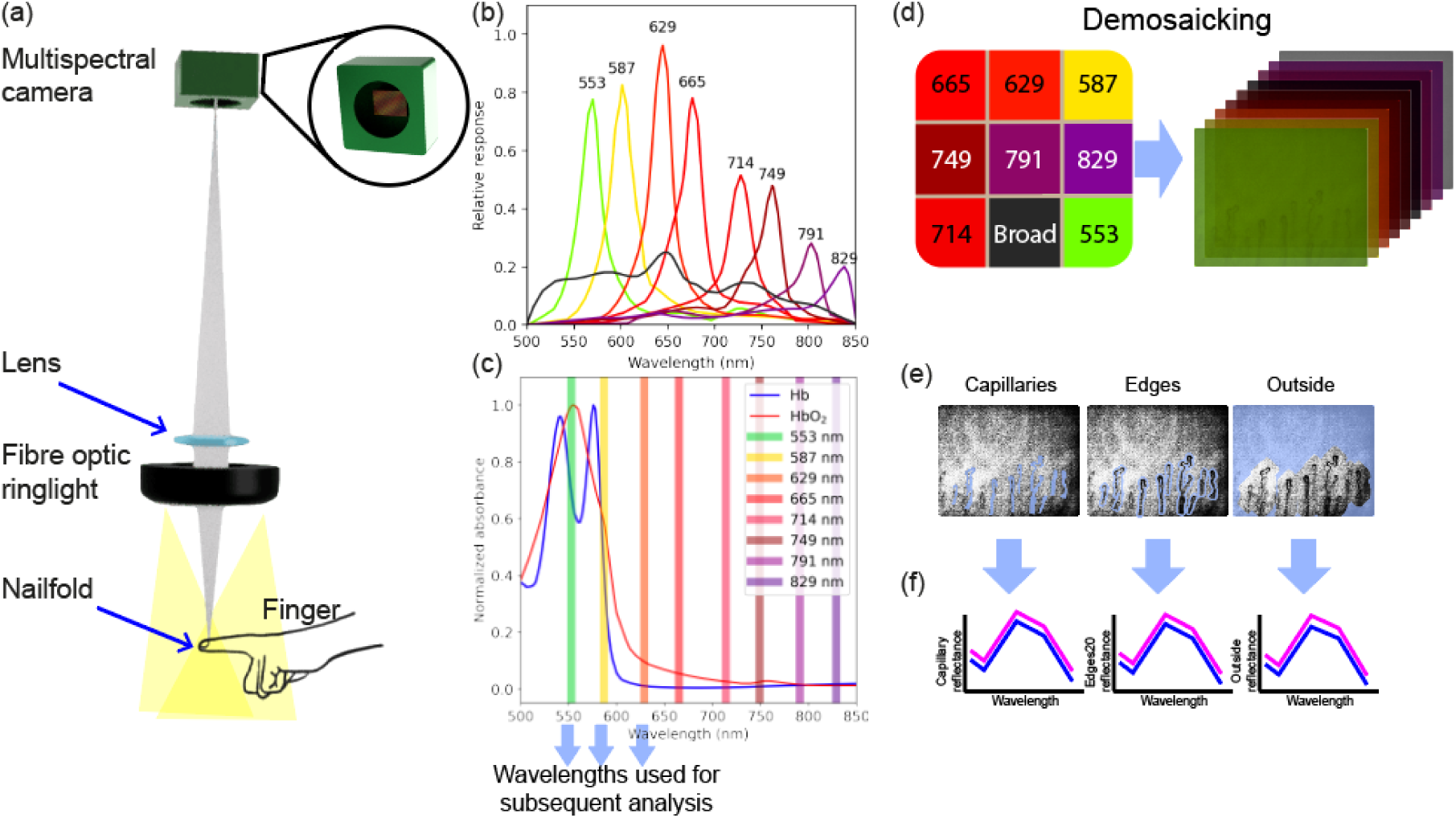
CAPoxy study imaging system and image processing overview. (a) An LED source illuminated the nailfold of the finger through a fibre optic ring light. Light reflected from the finger was magnified through a lens before being captured by the multispectral camera. (b) The spectral response of the MSI camera filters is shown for each of the 9 wavelength bands; 8 have a narrow bandwidth of ∼15 nm with centre wavelengths from 553 to 829 nm. The remaining band has a broadband filter with a passband from 500 to 850 nm (18). (c) Illustration of how the 9 wavelength bands sample the absorption spectra of oxy-(HbO_2_) and deoxy-haemoglobin (Hb). (d) The 9 filters (centre wavelengths noted) are arranged in a mosaic pattern on the MSI camera, such that each pixel is covered with one of the nine filters. The 3×3 pattern is repeated across the entire sensor, allowing 3D dataset to be collected in real-time (2D spatial information and 1D wavelength information). For ease of visualisation, the colour of the bands is depicted by the central wavelength, with variations of dark red / purples used for wavelengths greater than 720 nm, which is beyond our colour vision. Demosaicking converts each MSI camera image frame into a set of nine spectral images by identifying the pixels that correspond to each filter group and interpolating between them to recover a full image. (e) Three regions of interest (ROIs) were defined: capillaries, edges (within 20 pixels of the capillaries), and outside regions. (f) Spectra were extracted from each ROI.

Reflected light was magnified through a lens (f = 27 mm, Edmund Optics) before being captured by a multispectral camera (CMS-V, SILIOS). The system was mounted on three motorised stages (KMTS25E/M, Thorlabs) that allowed control of the system xyz position to easily focus and move along the nailfold. The multispectral camera collected images at nine wavelengths between 553 to 829 nm (Figure 1b, c). The modified system was characterised to ensure that any degradation in spatial resolution due to the use of the MSI camera (itself characterised previously (19,20)) would not impact the conclusions of the study (see Supplementary Methods: Imaging system characterisation) and was found to have a spatial resolution of 12 µm (Supplementary Figure 2a,b), which was suitable for application in capillaroscopy.

### Participants

This was a prospective single centre, cross-sectional, feasibility study to assess MSI in the nailfold. The study was approved by the Manchester Research Ethics Committee (REC reference number 22/LO/0537)’. and all participants gave written informed consent. Images were taken within December 2022 by a trained operator. Inclusion criteria for participants was over 18 years, able to provide written consent and, for patients with SSc, a confirmed diagnosis of SSc. The control group had no underlying vascular or skin conditions including no history of Rayanud’s phenomenon, a common precursor to SSc(9).

### Imaging protocol

Participants were acclimatised in a temperature-controlled room for 10 mins during which time the light source also equilibriated (Supplementary Figure 1b). Castor oil (chosen as colourless) was placed on each nailfold in turn, as imaged (to provide a smooth surface to improve visulaisation of capillaries within the skin); fingers were held in place with a grooved finger holder. Images were taken post participant acclimatisation in a darkened room to minimise illumination variations. Prior to each participant’s imaging, white images (exposure time of 46.4 ms) were obtained for each participant to allow calibration of the spectrum and spatial distribution of the LED light and dark images were obtained with no illumination to enable correction for background noise. Approximately 20 images were then captured across two nailfolds (both ring fingers), scanning to capture a wide field of view.

### Image and spectral processing

All image and spectral processing was performed using custom code written in Python and an extended description of the methodology can be found in the Supplementary Information (Supplementary Methods: Image processing and segmentation). Data, white and dark images were demosaicked (Figure 1d-f and Supplementary Figure 3) and then a correction procedure was performed (Supplementary Equation 1) to compensate for light source characteristics and dark noise properties. Corrected images were subjected to manual quality control to assess images for a) the presence of visible capillaries that could be segmented, and b) the absence of image artefacts occluding those capillary structures. Images that did not meet the quality control threshold (examples shown in Supplementary Figure 4) were excluded from the study.

Images that passed quality control were segmented into different tissue regions using a manual annotation so that the spectral properties of the different regions of the nailfold (capillaries, surrounding edges, and outside area) could be determined. Each visible capillary was manually outlined in Labelbox, an online software tool, on the image formed from the 553 nm spectral band on the MSI camera, which provided the best contrast between the the capillary and surrounding region. These outlines defined both the capillary mask and masks for surrounding regions according to established criteria (Supplementary Equation 2, 3). Masks were then applied to the images (Supplementary Figure 5). Subsequently, average signals (R) were computed in each region for each spectral band, generating arrays with nine spectral points per image (Supplementary Equation 2, 3).

The measured signals in this study are tissue reflectance. Reflectance refers to the amount of incident light reflected off the tissue surface. Reflectance in biological tissues is influenced by factors such as cellular composition, pigmentation, and structural organisation, which govern tissue absorption and scattering properties. To account for differences in overall reflectance between images and enable comparative evaluation of the entire dataset, the spectra were normalised to the measured intensity in the broad wavelength band, representing the integral of reflectance across the full wavelength range investigated. The normalised data were then examined on a per-participant basis, with average spectra plotted by region for each participant.

The normalised data were then used to train a random forest (RF) classifier (see Supplementary Methods: Random forest classification, Supplementary Equation 5, 6), with a range of classification metrics used to calculate performance. Briefly, random forest (RF) is an ensemble learning method used for classification that combines the outputs of multiple decisions trees to determine the optimum classification of tissue state (21). Individual decision trees have a series of root nodes, branches, and internal and leaf nodes to classify tissue (21). By using multiple decision tress often with different results, the decision trees then ‘vote’ such that the classification with the largest majority is then assigned to the sample. The RF classification was tested regionally, as well as by grouping all regions for the HC or patients with SSc (Supplementary Figure 6). Five metrics were used to analyse classification performance: classification accuracy, average cross-validation, precision, recall and F1-score (defined in Supplementary Table 1).

### Statistical analysis

All statistical analyses were performed using R Statistical Software (v4.2.3; R Core Team 2023). Linear mixed-effects models (LMMs) were analysed using the nlme R package (v3.1.162; (22)). The varExp and varIdent functions of the nlme R package were used to estimate respectively the residual variance at different wavelengths (primary analysis) and the region-specific residual variances (secondary analysis). The F-tests for linear combinations of LMMs’ parameters were performed using the anova function of the nlme R package. Best linear unbiased predictions (BLUPs) of spectral values and summary statistics at the participant level were obtained using the fitted function of the nlme R package. Outliers were identified using the robustbase R package (v0.99.0; (23)). Data visualization was performed using base R and the ggplot2 R package (v3.4.2; (24)).

In the primary analysis, LMMs were used to detect and estimate the dependence of the spectral bands on disease status and region. In the secondary analysis, LMMs were used to detect and estimate the dependence of the summary statistics AUC and maximum on disease status and region (see Supplementary Methods: Statistical analysis). For each subject, the summary statistics calculated from the average spectra were: the Area Under the Curve (AUC), the maximum over the eight spectral bands (bands illustrated in Figure 1b), along with a set of specific ratios between bands, chosen to highlight expected differences in the haemoglobin absorption spectra. The AUC was calculated using the trapezoidal rule. For the ratios, the following spectral bands (Figure 1c) were examined:

– The 553 nm band, which shows strong contrast between the capillaries and surrounding tissue but averages over peaks of both HbO_2_ and Hb, giving a mixed signal from both but expected to be somewhat weighted towards HbO_2_.
– The 587 nm band, where the absorption of HbO_2_ and Hb are relatively similar, so reflectance would be similar.
– The 629 nm band, where the absorption of HbO_2_ is substantially higher than Hb, so reflectance would be lower for increased oxygenation.

For the 553, 587, and 629 nm filters, due to the position of the center wavelengths, an increase in SO_2_ will result in an increase in reflectance. Despite this, there are differences because the rate of increase due to SO_2_ varies (Suppl. Figure 9). The rate that the 553 nm and 629 nm changes is similar. While the 587 nm changes less, about 50% less, so it can be approximated as isobestic, or impervious to changes in oxygenation. These differences can be used to understand the oxygenation map of the tissue.

We therefore examined the ratio 587 nm / 553 nm, where we expect changes to be more associated with changes in oxygenation of haemoglobin content, and 629 nm / 553 nm which is expected to be more affected by changes changes in haemoglobin content as the change due to oxygenation is approximately the same. Images were constructed by simply dividing each image by the other (after correction and normalisation). That is, each pixel of the 587 nm image was divided by the corresponding pixel of the 553 nm image. Additionally, simple denoising was performed to aid visualisation of capillary structures using a median filter (kernel=6).

## Results

### Example images show abnormal microvasculature in patients with SSc and diminishing contrast with increasing wavelength in both groups

The study enrolled 12 healthy controls (HC) and 10 patients with SSc; demographics in Table 1. Patients with SSc fulfilled the ACR/EULAR 2013 criteria for SSc (12). After quality control was performed on the dataset (see Methods), it was concluded that some images could not be processed; the following analysis was from 9 HCs and 7 patients with SSc. The data from three individuals from the HC group were fully excluded (one as it was a spectral outlier, and two as there were no images of sufficient quality to analyse, see supplemental data for examples). Likewise, three patients with SSc were excluded because there were no images of capillaries visible or sufficient contrast for analysis. Despite some challenges encountered during manual evaluation, such as out-of-focus images and low signal-to-noise ratio for the capillaries, image segmentation was successfully completed on 186 images from the 16 individuals.

**Table 1:** Summary of demographic data for the groups.

|  | **HC** | **SSc** |
| --- | --- | --- |
| Subjects, *n* (%) | 9 (56) | 7 (43) |
| Age, median (IQR) years | 43 (34 -55) | 54 (49- 65) |
| Sex, female (%) | 7 (77) | 7 (100) |
| Duration of RP, median (IQR) years | - | 20 (10-24) |
| Duration since first non RP symptom, median (IQR) years | - | 10 (6-21) |
| Severe digital ischaemia*, *n* | - | 2 |
| Digital ulceration on the 12 months, *n* | - | 2 |
| Digital pitting, *n* | - | 2 |
| Number of RP episodes in the last week, *n* (IQR) | - | 21 (18-35) |
| RCS, *n* (IQR) | - | 5 (3-7) |
| Images taken, *n* | 168 | 129 |
| Images used, *n* | 91 | 75 |

The images of the nailfold were first examined qualitatively at each individual imaging wavelength and showed varying degrees of capillary visibility (Figure 2), as would be expected based on the relative haemoglobin concentration and absorption spectra (Figure 1c). The 553 nm band images showed the highest contrast and capillaries are also easily visible in the 587 nm band and broadband images. At higher wavelengths (above 629 nm), the images do not have high contrast due to the diminishing absorption of light by haemoglobin (Figure 1c), however, the capillaries can still be observed albeit at low contrast.

**Figure 2:**
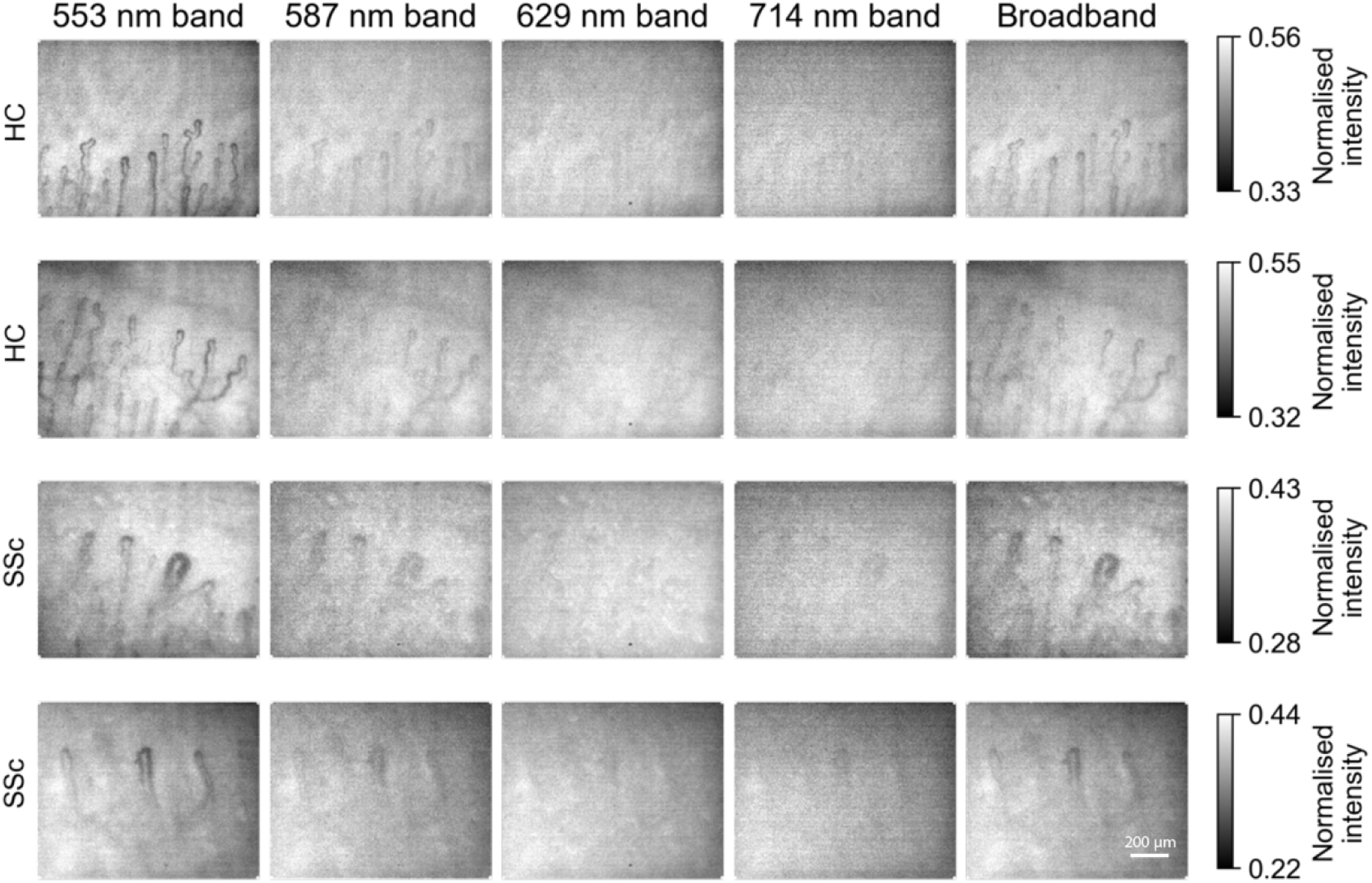
Example images of two individuals from the HC group and two patients with SSc. Data are presented at a subset of five different bands: 553 nm, 587 nm, 629 nm, 714 nm, and broadband, to demonstrate the differences in imaging contrast according to wavelength. The contrast for the images has been scaled for comparison purposes and is shown on the right side of each row.

### Initial data exploration suggests subtle spectral differences between healthy controls and patients with SSc

The average spectra from each imaging region (capillaries, surrounding edges and outside) were first investigated by comparing the average spectra from individuals and across the HC and SSc groups (Supplementary Figure 7).

Summary spectra plotted by region for the HCs (Figure 3a,c) and patients with SSc (Figure 3b,c), show that the highest reflectance occurs at 629 and 665 nm, likely due to strong haemoglobin absorption outside of that region. Average spectra differed between participant groups and spatial regions. Adjusted for different baseline values at 553 nm, patients with SSc had slightly higher reflectance values for all the bands above 553 nm, though this finding was not statistically significant (p=0.22), likely due to the small sample size and high variance in the data.

**Figure 3:**
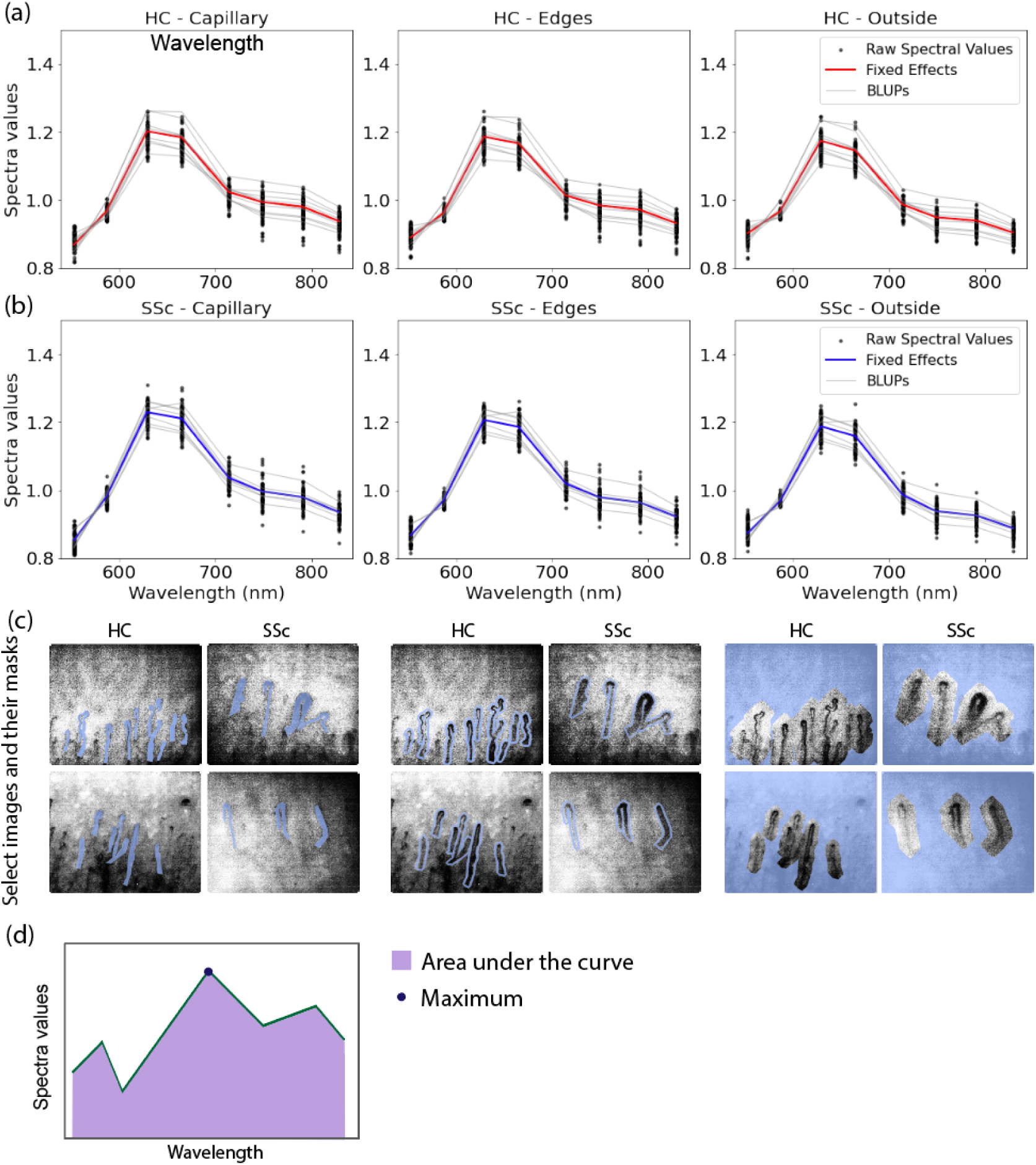
Comparison of spectral properties for the two groups. (a) healthy control group and (b) patients with SSc. Dots are the raw spectral values for each participant. Red and blue lines show fixed effects attributable to wavelength, region and disease status. Grey lines show the best linear unbiased predictions (BLUPs) of the participant-specific random effects. (c) Example images and the masks for each region are illustrated – left, capillary; middle, edges; right, outside. (d) Illustration of the AUC and maximum calcualted from the spectra.

In an exploratory analysis, the difference between HC and SSc groups for all bands above 553 nm was smaller in the regions outside capillaries (p<0.001), as would be expected given that the region outside of the capillaries is less likely to contain light-absorbing features. The outside region showed two other effects: lower absolute reflectance values for all bands above 553 nm (p<0.001), and increased baseline reflectance values above 553 nm (p<0.001). Both effects were common to HC and SSc groups. Finally, it was noted that the variability of the spectral values (single points in Figure 3a and b) was different between wavelength bands (homogeneity of residual variance across the wavelengths was tested using the likelihood ratio test with restricted likelihood estimator: p<0.001).

### Statistical analysis reveals spectral differences between HC and SSc groups that underpin classification

Although classification was not the specific purpose of the study, a random forest (RF) classifier was tested (Supplementary Figure 6) to examine whether the spectral features could provide a basis for classification given the subtle spectral changes observed. The highest classification accuracy found was 0.84; this corresponds also to the highest average cross validation, precision, recall, and F1-scores. Classification based on spectra from different regions was also examined and the highest average classification accuracy (0.82) also occurs when employing RF on the whole dataset (Supplementary Table 2). The corresponding individual accuracies are all similar: 0.87, 0.82, and 0.79 for the capillary, edges, and outside region, respectively (Supplementary Table 3). Other classification techniques were examined (k-nearest neighbors and support vector machine), but had lower classification accuracies.

Summary statistics support the results of the primary spectral analysis, which showed subtle differences between the groups, and subsequent exploratory classification analyses, which showed a reasonable accuracy of classification based on those differences. SSc subjects have slightly higher AUC and maximum values than HC subjects (Figure 4; p_AUC_=0.48 and p_max_=0.19). Importantly, the difference between HC and SSc groups was specifically in the capillary region, as the AUC and maximum values (p_AUC_<0.001 and p_max_<0.001), which would be expected since the capillary regions primarily contain the haemoglobin in red blood cells that dominates the spectral characteristics. For both groups, AUC and maximum statistics have lower values in the outside regions (region effect, p_AUC_<0.001 and p_max_<0.001). These changes can be visualised in individual images of the capillaries (Figure 5 and Supplementary Figure 8).

**Figure 4:**
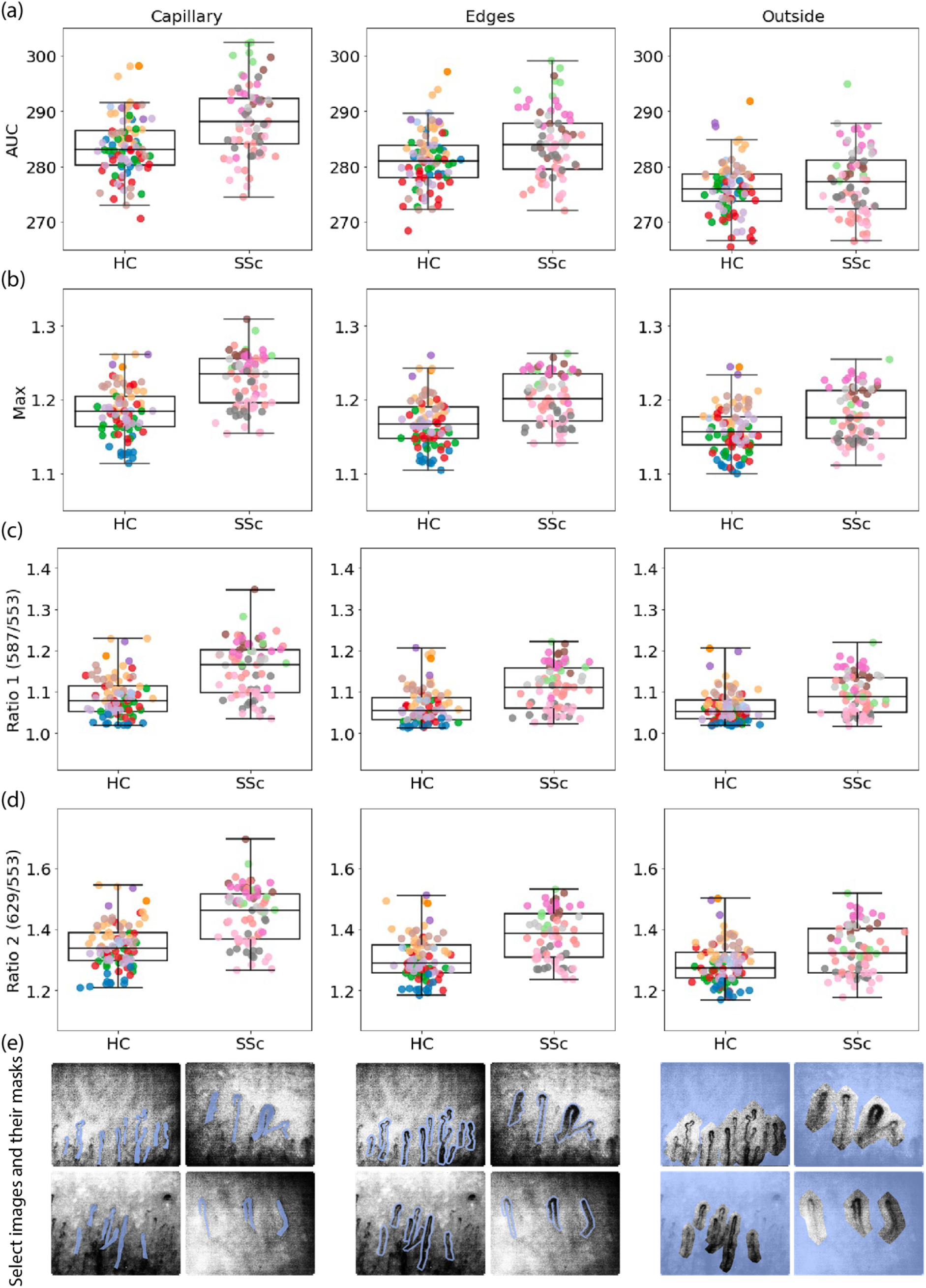
Comparison of differences in summary statistics between HC and patients with SSc. (a) AUC, (b) maximum, (c) ratio 587 / 553 nm, and (d) ratio 629 / 553 nm. Each participant is denoted by a different colour to illustrate the interparticipant variation and the effect of disease. Their distribution is illustrated by box-plots. (e) Example images and their masks are illustrated.

**Figure 5.**
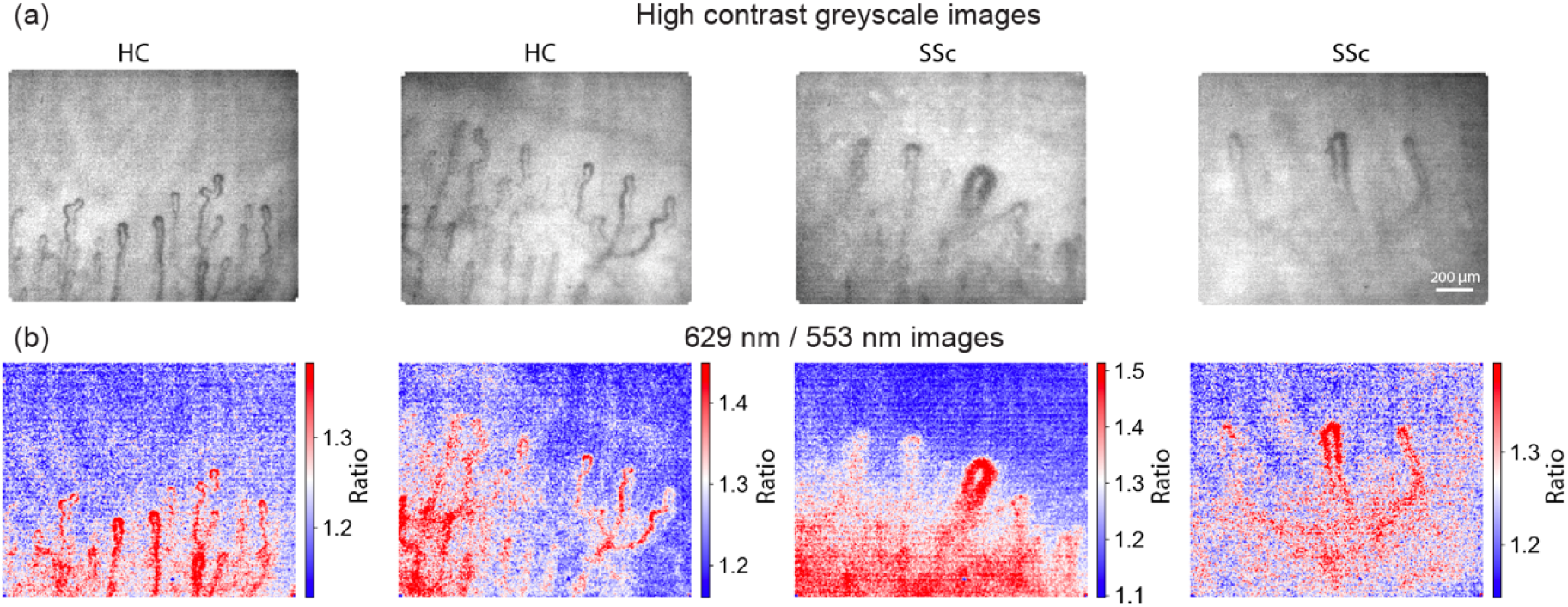
Exemplar false-colour ratiometric images demonstrated oxygenation weighted images. (a) the greyscale image taken at 553 nm, (b) the ratio of the 629 nm / 553 nm images after denoising.

For all ratios examined (Figure 4c), a region effect was detecting showing that the behaviour of the capillary region is different from all other regions, with a trend to statistical significance. The 587 / 553 ratio increased from 1.109 ± 0.045 (Mean ± 95% CI) across the HC group to 1.167 ± 0.052 across the patients with SSc. The 629 / 553 ratio increased from 1.375 ± 0.069 across the HC group to 1.462 ± 0.071 across the patients with SSc. The spectral findings support these ratiometric changes. A qualitative visualisation of the ratios as images was also made (Figure 5), showing the potential to enhance grayscale images (Figure 5a) with oxygenation-weighted contrast (Figure 5b, Supplementary Figure 8) for visual inspection. While it is difficult to see the effects on individual patients, the overall ratios indicate that the capillaries and surrounding tissue are less oxygenated in the SSc group than in the HC group.

## Discussion

This feasibility study, investigating the application of MSI in nailfold capillaroscopy, demonstrated that MSI allows visualisation of the capillaries in most participants across the HC group and patients with SSc, enabling the analysis of spectral features. The capillaroscopy spectra obtained were found to discriminate between particpant groups but differences were found to be subtle; normalisation of the spectra was applied to aid interpretation. Regional analysis was performed to obtain additional insights into the spectral differences within the capillaries themselves, the surrounding edges, and outside regions.

While subtle, spectral differences could be exploited for image classification, with a RF classifier showing accuracy of 84% overall, combining data from all regions, and 82% when looking at individual regions. RF was chosen for this purpose as it is able to handle large noisy datasets and nonlinear relationships between the different data groups.

All summary statistics reinforced the ability to discriminate between groups, with increased AUC and maximum of the respective regions analysed seen in patients with SSc, indicative of the differences in the overall reflectance between groups; the HC group generally showed increased reflectance. During manual annotation of these images, it was observed that patients with SSc often have more haemorrhaging of blood outside of the capillary structure and larger capillaries, which could explain why they display more absorption in the nail bed and hence decreased reflectance. The HC group also showed elevated oxygenation weighted ratios, which would be expected given the decreased oxygenation expected with the evolution of SSc. The ratio of the 587 nm to 553 nm for the SSc patients was 5% higher than the HC group.

Although the spectral differences in HC and SSc tissues are subtle, they can be distinguished, and MSI coupled with machine learning algorithms could potentially be an effective tool for the automated evaluation of SSc when combined with the existing morphological features.

Regional analysis could then potentially contribute to a more comprehensive understanding of the microvascular changes in SSc and aid in targeted diagnostic assessments.

Despite these promising findings, this was a feasibility study with a broad eligibility criteria and small sample size, meaning that it is difficult to generalise findings and a confirmatory study would be needed before making general conclusions. The small sample size meant that the level of variation in the data was large in comparison to the spectral differences observed between HC and patients with SSc. Furthermore, the observed differences between the HC group and the patients with SSc could be heavily confound by other factors heterogeneously distributed between the two groups, such as age and sex of participants, or differences in skin tone that could influence the results. These would need to be well controlled in any confirmatory study.

In addition to these study limitations, there are also some technical limitations that could be addressed. The imaging system was operated with a white light LED, which lacked sufficient illumination power to achieve good signal-to-noise ratio in the longer wavelengths greater than 714 nm. Subsequent ongoing work has substituted the LED with a halogen light source to improve illumination at these wavelengths and better understand additional benefits that could be derived from including them in the MSI system. Future work imaging nailfold capillaries should also encompass the shorter wavelengths between 400 to 550 nm, as they are expected to demonstrate greatest contrast between capillaries and the surrounding tissue due to greater haemoglobin absorption in this range. Adding further blue wavelengths might also enable linear spectral unmixing of absolute oxygenation values in the nailfold capillaries(15,17), rather than the oxygenation-weighted ratios used here. However, this may be impacted by skin tone to a greater extent.

With future improvements in the spectral targeting and classification, the clinical implications of MSI in nailfold capillaroscopy are potentially far-reaching. It would also be interesting to combine the measured spectral information with standard morphological information that is typically used in clinical interpretation of capillaroscopy images. Adding functional information with MSI to capillaroscopy has the potential to better characterise microvascular dysfunction, including blood flow characteristics and oxygenation, which could be relevant patients with SSc and also across a wide range of diseases, such as diabetes, hypertension, and connective tissue disorders.

The feasibility study undertaken here demonstrates that it may be possible to use MSI to measure oxygenation in the nailfold capillaries of patients with SSc. A further confirmatory study is required to demonstrate the results of this feasiblity study.

## Disclosure

The authors declare no conflicts of interest.

## Author contribution

AM and SEB supervised the study. MT-W modified the optical system and performed calibration measurements. Images were obtained by JM and IK. MT-W analysed images and data discussed in this manuscript. MB, GD, and AM provided advice on the data acquisition and computational analysis. MT-W and LP performed statistical analysis on the data set. AM designed the protocol, had oversight of the imaging study and had oversight of image analysis. AM and SW prepared the research ethics submission. MT-W drafted the manuscript and supplementary information and prepared all the figures and data sets. All authors revised the manuscript.

## Funding

MT-W would like to acknowledge the financial support of the General Sir John Monash Foundation, the Cambridge Trust International Scholarship, and the Cavendish Laboratory’s AW Scott Scholarship. SEB was funded by Cancer Research UK under grant number C9545/A29580 and the UKRI Engineering and Physical Sciences Research Council under grant numbers EP/X037770/1 and EP/R003599/1. This research is co-funded by the National Institute for Health and Care Research (NIHR) Manchester Biomedical Research Centre (BRC) (NIHR203308). The views expressed are those of the author(s) and not necessarily those of the NIHR or the Department of Health and Social Care.

## Data Availability

The data underlying this article will be shared on reasonable request to the corresponding author.

## Notes

### Competing Interest Statement

The authors have declared no competing interest.

### Clinical Trial

NA

### Funding Statement

Yes

### Author Declarations

The study was approved by the Manchester Research Ethics Committee (REC 22/LO/0537) and all participants gave written informed consent.

